# Pathways to optimize a pediatric telemedicine and medication delivery service: A multi-level qualitative study in Haiti

**DOI:** 10.1101/2025.03.26.25324692

**Authors:** Frantz Emmanuel Garilus, Kerven Cassion, Youseline Cajusma, Katelyn E. Flaherty, Jude Ronald Beausejour, Lerby Exantus, Valery M. Beau de Rochars, Chantale Baril, Torben K. Becker, Rochelle K. Rosen, Eric J. Nelson, Molly B. Klarman

## Abstract

**Background:** While telemedicine has become an established component of healthcare delivery globally, challenges to scaling emerging initiatives persist across multiple levels. Over the last 5 years, our team has developed a pediatric telemedicine and medication delivery service (TMDS) in Haiti that integrates clinical guidance with rapid access to medications. Building on successful proof-of-concept studies, we are now well positioned to characterize both general challenges to telemedicine scale-up and those unique to the TMDS model.

**Methods:** In this study, we conducted focus group discussions and administered written questionaries to TMDS staff, including physicians, nurses, and medication delivery drivers. Using framework matrix analysis we identified key challenges and opportunities associated with the TMDS model.

**Results:** Areas for improvement related to obtaining quality information from virtual exams, the reliability of technology and communication infrastructure, conditions necessary for effective in-person exams, the limited scope of the TMDS workflow and clinical resources, and uncertainty surrounding long-term sustainability. These insights informed the development of targeted action items categorized into three domains: conceptual, physical and mission-oriented.

**Conclusion:** The findings will guide our internal scale-up strategy and may offer guidance to similar telemedicine initiatives.

## INTRODUCTION

In the wake of the COVID-19 pandemic, telemedicine became a central fixture in healthcare infrastructure [1]. A post-pandemic analysis of the utilization of telehealth and telemedicine services found at least 77 countries had adopted the approach [2]. Telemedicine, generally defined as ‘*the use of technology to provide healthcare services between parties that are geographically separated’* [3, 4], is rapidly diversifying in scope [5]. One categorization of telemedicine uses five application types: tele-consultation, tele-diagnosis, tele-ambulance, tele-monitoring, and digital self-management. The perspective considers six additional descriptive components: personnel, target population, setting, technology, data provision and intended outcome [6]. Permutations of these components demonstrate the broad range of possible telemedicine applications.

The benefits of telemedicine vary depending on the specific application or use case. One use case is a type of synchronous (real-time) teleconsultation between provider and patient known as ‘triage and treat’. In this model, patients are remotely screened to determine the appropriate location and method of treatment [7, 8]. In low-resource settings, the triage and treat approach can increase access to specialists, address shortages of healthcare workers, and reduce transportation related barriers, including time and cost [9–11]. With these goals in mind, our team developed a pilot pediatric telemedicine and medication delivery service (TMDS) that has been deployed in Haiti and Ghana [12, 13]. In the TMDS model, patients are first triaged and then assessed through a virtual exam, followed by the delivery of medications directly to their homes. To the best of our knowledge, there are few other (if any) affordable, high-quality, evidence-based, pediatric TMDS interventions deployed in resource-limited settings. As with many aspects of telemedicine’s evolution, this is likely to change.

Scaling novel healthcare interventions, such as telemedicine, is challenging, particularly in low-resource settings [14–16]. A broadly used definition of ‘scaling up’ a health innovation is, ‘*deliberate efforts to increase the impact of successfully tested health innovations, so as to benefit more people and to foster policy and program development on a lasting basis’* [17]. Scaling is unlikely to occur organically, even with the most promising innovations [14]. Barriers to scale can be numerous and present across various implementation domains[18].

One common approach to identify these barriers is through frameworks such as the Consolidated Framework for Implementation Research (CFIR) [19]. CFIR covers five domains: innovation characteristics, outer setting, inner setting, characteristics of individuals, and implementation process. These domains can help assess barriers to implementing telemedicine interventions. Within the ‘innovation domain’, issues with poor interoperability are common [20, 21]. Within the ‘outer setting’ many low-resource countries lack nationwide policies and regulatory standards for telemedicine [22–24]. Within the ‘inner setting’ challenges to obtain and maintain information and communication technology infrastructure (ICT) are often cited [23, 25]. Within the ‘individuals’ domain both providers and patients have expressed hesitancy to accept telemedicine [26, 27]. Within the ‘implementation process’ domain many telemedicine programs were rapidly implemented in response to COVID-19 and did not undergo the necessary planning stages to support sustained success [28]. In addition to identifying barriers, stakeholders are also committed to recognizing the facilitators that support successful, and scalable implementation.

Qualitative methods play an integral role in implementation science, allowing implementors to systematically identify barriers and facilitators to successful initiatives. The methods allow for systematic and efficient description of what is happening and why it is happening [29]. Proctor et al. proposed eight outcome measures for implementation research, of which five recommend qualitative research methods as the primary measurement tool [30]. Similar to quantitative research, the research question leads decisions on which data collection and analysis methods are most appropriate [31]. Among various approaches, framework matrix analysis is especially effective for producing rapid, streamlined summaries that support deductive learning and can be easily shared between diverse stakeholders [32].

With the objective of prioritizing early access to pre-emergent healthcare to prevent the need for more costly and complex emergency care, our team developed a nighttime pediatric telemedicine and medication delivery service (TMDS) in Haiti. The service has evolved within the Improving Nighttime Access to Care and Treatment-Haiti (INACT-H) study series. The TMDS model was designed to overcome the healthcare access barriers of distance and nighttime [33]. From 2019 to 2021, we validated this innovative healthcare delivery model, finding that it was clinically safe, feasible to implement, and supported by reliable clinical resources [12, 34]. From 2021 to 2023, we focused on positioning the TMDS to scale by modifying the workflow. The resulting scalable model maintained strong clinical and operational outcomes [35]. A cost-effectiveness analysis confirmed that a scaled TMDS was a cost-effective alternative to hospital emergency care for pre-emergency pediatric conditions at night in Haiti [36]. In this context, we sought to qualitatively identify challenges and opportunities that could improve both cognitive and physical factors to optimize the TMDS for large-scale implementation.

## METHODS

### Study setting

A TMDS called MotoMeds was launched in the semi-urban and rural region of Gressier, Haiti in September 2019. The workflow of the TMDS is as follows (Figure 1): a parent calls the call center regarding their sick child. A TMDS nurse (registered nurse or family nurse practitioner) collects demographic and clinical findings from the parent to complete a virtual exam by phone. Nurses use TMDS specific clinical resources to generate an assessment and treatment plan that includes illness severity, disposition, medication/fluid recommendations, and advice for follow-up care. Illnesses triaged as severe are referred directly to emergency services. Among children who reside within a delivery zone, those with higher clinical uncertainty are offered an in-person household exam with delivery and patients with low clinical uncertainty are offered a delivery of medications/fluids. Children who reside outside of a delivery zone are provided with guidance alone. All children receive a follow-up call at 10-days and patients with severe illness also receive a call at 24-hours. On-call physicians are available by phone to consult on cases beyond the scope of the TMDS clinical resources.

**Figure 1.**
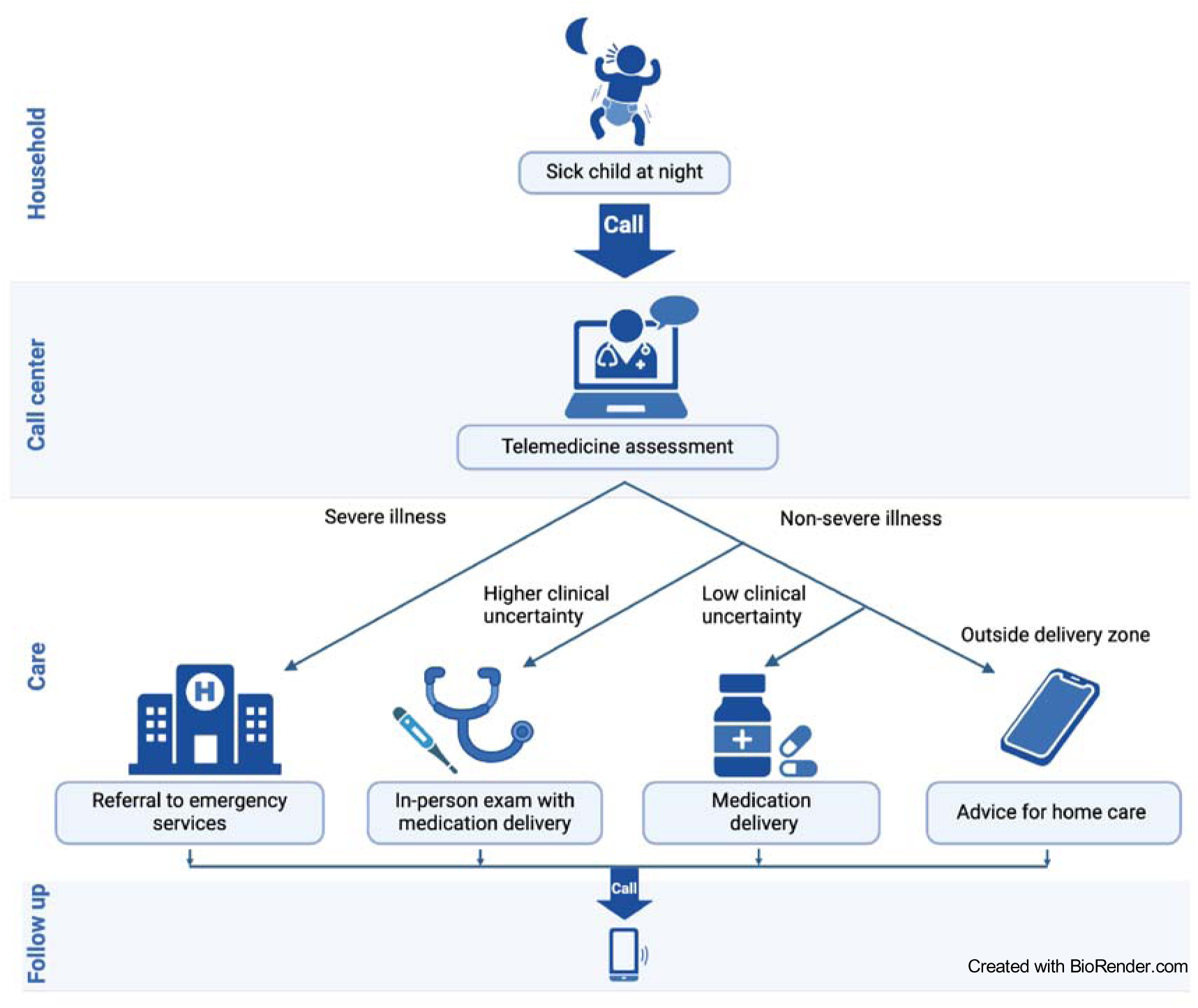
General workflow of a telemedicine and medication delivery service (TMDS). The parent of a sick child contacts the call center. A TMDS nurse receives the call and conducts a virtual exam by asking the parent targeted questions about the child and their symptoms. The nurse references clinical resources to create a treatment plan that includes disposition, medications/fluids, and follow-up recommendations. Families are contacted by phone at 24 hours and/or 10 days. A version of this figure was published previously[35].

In October 2021, a second delivery zone was established in the urban center of Les Cayes, located 175 Km west of Gressier. The workflow of this delivery zone differs in that treatment plans generated at the central call center in Gressier are relayed to on-call nurses in Les Cayes who either dispatch motorcycle delivery drivers to transport medications/fluids or conduct in-person household exams. Once proofs of concept were established through three years of evaluation, iteration, and growth, our team pivoted towards positioning the TMDS for scale. We sought qualitative feedback from TMDS staff as a critical component of this process. The research team consisted of EJN, the PI who is a pediatrician and scientist with qualitative research experience [32, 37], MBK, the study director who is an international public health specialist, and two bilingual Haitian American facilitators, FG and KC. Each team member was coached in qualitative methods by RKR, an expert in qualitative methods including matrix analyses applied in global health settings challenged by technological and situational limitations [32, 38].

### Study design

#### Focus groups

EJN, MBK, FG, and KG developed a written agenda (Additional File 1) to guide focus group discussions for each of four types of TMDS staff/participants. Participant types included: i) call center nurses, who conduct virtual exams at the call center and relay treatment plans, ii) delivery nurses, who receive and enact virtual treatment plans by dispatching a motorcycle delivery driver or conducting in-person household exams, iii) on-call physicians, who provide consultations by phone, and iv) motorcycle delivery drivers, who transport nurses and deliver medications/fluids to households. Most questions and discussion prompts were specific to participant type, but a subset was shared across all participants. Topic areas were identified *a priori* as either factors that could enable or impede scaling (clinical, technical, and logistical challenges and solutions) or that qualitatively assess impact (family expectations and satisfaction, and value to TMDS staff and community). The agenda was translated to Haitian Creole by FG and KC. MBK reviewed the agenda questions with a TMDS staff member to ensure the questions were relevant to the staff experience.

In June and July 2022, FG and KC conducted nine focus group discussions via video conferencing software (Zoom) with facilitators based in the United States and participants located in Haiti. All active staff members were offered participation and were placed in groups of 3-4 based on type and location. At the beginning of each session a facilitator read a waiver of documentation of written consent statement to each person individually. After expressing verbal agreement, participants were given a study ID. The video sessions were recorded.

#### Questionnaires

Two versions of a nurse questionnaire were developed by MBK with 26 short answer questions for call center nurses and 15 for delivery nurses (Additional File 2). The questionnaire focused on identifying ways to improve the clinical resources and procedures that guide virtual and in-person exams. Given the growing interest in clinical decision support (CDS) tools, particularly digital versions of World Health Organization guidelines, the questionnaire also assessed interest in integrating electronic clinical decision support into the TMDS workflow.

### Data analysis

#### Data preparation

FG and KC translated (Haitian Kreyol to English) and transcribed the focus group video recordings in a single step. One facilitator completed a focus group in its entirety and the other reviewed it for accuracy, which included confirming the translation and correct identification of the speakers. Discrepancies were discussed between the facilitators and if necessary, a third team member (MBK) was consulted. Questionnaire responses were translated into English using the same procedure applied to the video recordings.

#### Matrix analysis

Four matrices were deductively constructed in Microsoft Excel. The column headers were drawn from the focus group agenda and nurse questionnaire and the rows corresponded to each participant. To populate the matrices, one facilitator read a transcript or questionnaire question by question and summarized the response by each participant in the corresponding cell, separating unique ideas with bullet points.

The opposite facilitator reviewed the transcript or questionnaire and the matrix for completeness and a third team member reviewed the matrices for clarity. At the bottom of the matrix 4 additional ‘summary rows’ were created; in the 1^st^ row, MBK summarized the responses from all participants for each question, in the 2^nd^, 3^rd^ and 4^th^ rows FG, KC, and MBK suggested possible action items to address ideas presented by participants. Through Zoom the four-member team reviewed the ‘summary row’ cells question by question and through discussion established a ‘consensus of summary’ and a ‘consensus of action items’ which were entered into additional rows of the matrix spreadsheets. Common domains relating to barriers and facilitators for scaling, and potential action items to improve the service were identified. FG and KC reviewed focus group transcripts and identified quotes that supported the common domains.

## RESULTS

### Study overview

All active staff (14 nurses, 13 drivers, 3 on-call physicians) agreed to participate in one of nine focus group discussions (Table 1). In addition, 14 nurses completed a written questionnaire. Participant input was captured within five domains: 1) Virtual exams at the call center, 2) In-person exams at the household, 3) Clinical resources, 4) Communication, and 5) Expectations and impact.

**Table 1.**
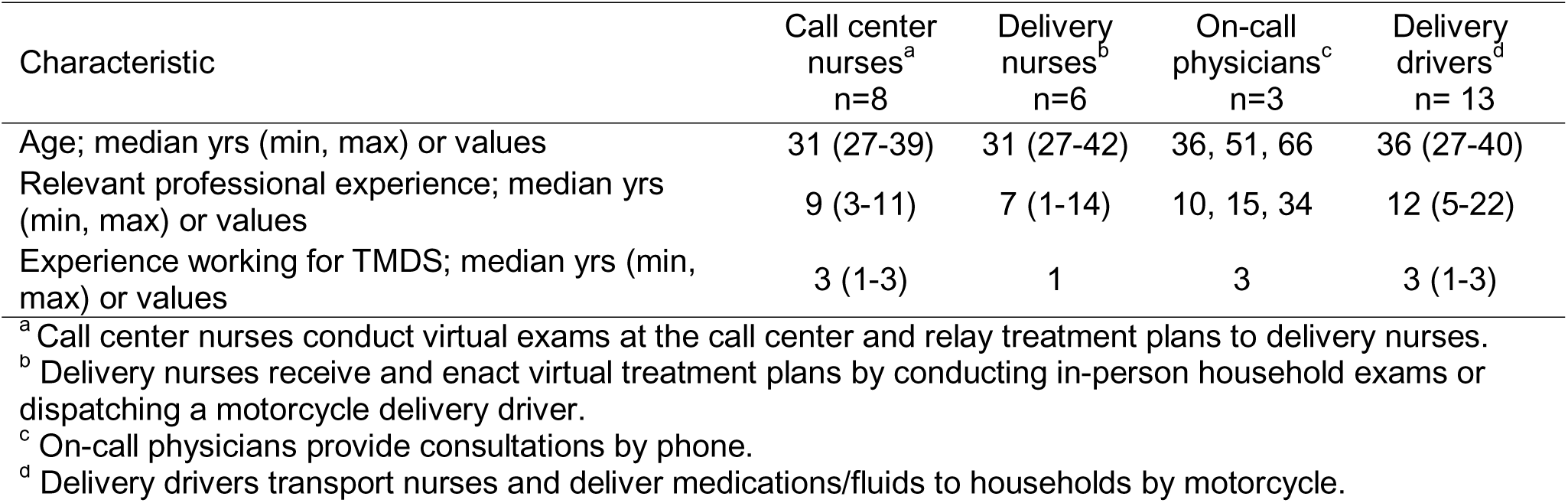
TMDS staff participant characteristics.

### Virtual exams

When discussing how nurses perceive the information parents relay over the phone, nurses stated they felt confident in the information families shared when responses were provided without hesitation, consistent over time, and aligned with other responses. They doubted the accuracy of the information when they had to repeat a question multiple times, and when they received inconsistent or implausible responses. One nurse explained, *“…sometimes we ask questions, and they* [callers] *are bouncing between ‘yes’ or ‘no’. This makes it difficult for us to have confidence in the information we are receiving. What also happens, when we do not have confidence, we mark ‘not confident’* [on the case report form]*. For example, if you have asked your first question and the answer is ‘yes’, and for things to correlate, the answer to the second question must also be ‘yes’, but the parents say ‘no’. Thus, there is a lack of correlation, and this makes us lack confidence in the information we are receiving.”* In these situations, nurses stated that they attempted to improve the data quality by rewording questions and explaining the importance of providing accurate responses. If they remained unsure about a particular response, they marked ‘not confident’ on the case report form and that response did not factor into the treatment plan. Nurses reported that specific components of the virtual exam were particularly challenging to complete. They felt that families were often resistant to identifying a chief complaint among multiple complaints and struggled to measure the respiratory and heart rates. Multiple nurses noted that questions related to respiratory problems (e.g., stridor, retractions) were difficult for parents to understand and often required repeat explanations and/or exploratory follow-up questions that were not part of the structured virtual exam.

### Call center characteristics

When reflecting on the work environment, nurses attributed the effective operation of the call center to the consistent availability of necessary materials and strong teamwork among the nursing staff. In contrast, unreliable internet and cellular signal were obstacles to performing call center responsibilities. Although technology is integral to the TMDS workflow, frequent changes to user-interfaces, such as laptop password expirations and software updates, along with unfamiliar information technology (IT) issues were cited as impeding call center operations. One nurse commented, *“…problems we have are that in the [software] applications, you connect to your account in a specific way and in a moment it changes. This means the nurse must have knowledge in informatics. If all the nurses did not have knowledge in informatics, this would be a big problem for us. I can come today to sign into my account to replace the nurse that was here last night, and the way I am used to signing in, I can’t sign in that way anymore.”* IT management strategies involved relying on contingencies (secondary internet carrier) and contacting the Study Director.

### In-person exams

During in-person exams nurses faced both technical and clinical challenges. Technical challenges included inadequate lighting and space, a lack of flat surfaces to place a scale to weigh patients, and missing materials due to failure to verify supply levels before departure. Clinical challenges included difficulty obtaining vital signs from young children, parental hesitancy to obtain temperature rectally, and a feeling of incompetency when the TMDS clinical resources or formulary did not cover a patient’s situation or when contacting the on-call physician in the presence of the parents. One nurse said, *“A difficulty we face performing in-person exams is when I encounter a complex case and I need to call the on-call physician in front of the family, they might think I am incompetent”*. Despite these challenges, nurses valued several aspects of the in-person exam, including obtaining vital signs, exploring the patient’s medical history, and administering the first dose of medication.

### Household logistics

Nurses commonly reported challenges reaching participant households, which they attributed to the difficulties of nighttime travel by motorcycle, poor road quality, remote household locations, and inability to connect with the parent by phone upon arrival. Nurses reported spending substantial time explaining the consent process, educating parents on how to administer medications, and discussing the rationale for the TMDS guidelines (e.g., why it only serves children ≤10 years or only covers specific illnesses). A mismatch between the parameters of the TMDS and parental expectations also required dialogue. Nurses described parents wanting certain medications that were not indicated (e.g., an antibiotic for a cough), requesting in-person exams for additional unenrolled children at the household, and encountering a different clinical situation than what was relayed during the virtual exam. One nurse commented, “*The problem I face is that when we reach their house, they tell us that they have other children that are sick and if we could see them also. We tell the parent to call the call center first, but they don’t agree. They want us to examine all their children, although they did not originally call for that. Another issue that we face is that the problem the parent calls for is not fully accurate. When we get there, we see that the child has more problems than they [originally] told us*.” The TMDS clinical guidelines recommended that patients with severe illness seek emergency level care. However, parents of children with severe conditions frequently reported being unable to do so due to financial constraints. In these situations, families still expected to receive medications, despite delivery nurses not being equipped to manage severe conditions. One nurse shared, “*Another problem is that MotoMeds does not treat every problem, some problems that a child may have, they are in the red [severe] category, you must send them to the hospital. Sometimes when we get to the house, with the state of the child, we can’t give them anything and we must send them to the hospital. But the parents don’t have the means and they are not happy that we didn’t give their child any medication. The parents feel as though they didn’t get any service because of this*.”

### Clinical resources

The nurses generally expressed satisfaction with the clinical resources, stating that they rarely encountered clinical situations beyond their capacity to manage. When faced with situations outside the scope of the clinical resources, they contacted an on-call physician, as the workflow intended. However, nurses did note occasional challenges related to medication dosing. They expressed concern about the inability to divide pills evenly, or at all, if the medication was only available in capsule form. Further, nurses were concerned that the estimated weights they used to perform dosing calculations could misrepresent patients’ actual weights in the context of undernutrition.

The role of the on-call physician was evaluated from both the nurses’ and physicians’ perspectives. Nurses suggested that physicians may not be fully aware of the constraints they face, such as being limited to the medications listed in the formulary. In some instances, physicians recommended medications not included in the formulary, even when equally effective alternatives were available within it. Despite the known limitations of telemedicine, physicians reported receiving sufficient case details to offer consultation. However, they noted that having a mechanism to receive photos or videos would be beneficial. It was broadly acknowledged that the on-call physician role is crucial but could be improved by developing a formal tool for gathering and relaying case details, as well as providing physicians with feedback on the outcome of each case they consult. Additionally, despite feeling connected to project leadership through WhatsApp groups and weekly email updates, on-call physicians felt disconnected to the broader TMDS team.

In response to the possibility of incorporating an electronic clinical decision support (eCDS) tool into the TMDS workflow, all nurses supported the adoption of this approach. They believed it would increase efficiency, conserve resources, and improve the quality of clinical records/data collection. However, it was noted that the tool should not depend on a stable internet connection.

### Communication

Given the critical importance of effective communication across the multiple channels within the TMDS workflow, all participants were asked about their experience communicating with staff and families. Call center nurses consistently cited an unstable cellphone network as their primary barrier to communication with delivery nurses and drivers. Similarly, delivery drivers also identified the unstable cellphone network as their main obstacle to communicating with nurses, other drivers, and families. The inability to reliably reach families by phone posed challenges to locating homes, particularly in a setting without formal addresses. In such situations, drivers relied on regional landmarks obtained during the virtual exams, along with their own local knowledge to find households. Drivers suggested the use of radios or higher quality cellphones to improve communication. They also recommended that nurses prioritize collecting formal names of landmarks and individuals instead of nicknames. Call center nurses, in turn, suggested several improvements, including ensuring that delivery nurses and drivers are prepared at the beginning of each shift, having checked that phones are charged and have sufficient cell minutes and data. The delivery nurses proposed working from a dedicated office rather than from home, and additional training with technology.

### Expectations

Through both virtual and in-person interactions with families, nurses and drivers identified several expectations families had for the TDMS. Nurses reported that families generally expected and received reliable, timely care, and quality medication. However, nurses felt that families also held expectations for services beyond the program’s scope, including care for patients older than 10 years, specific medications (e.g., cough syrup), service during daytime hours, and financial assistance to access emergency care when referred. As one nurse explained, *“Sometimes when a parent calls with a problem that surpasses our limits, we ask them to go to the hospital. The parent usually expects that we would do something for them before they go to the hospital. But if we send them to the hospital, there is no visit or medication that is sent to that house*.” Drivers reported observing expectations similar to those noted by nurses but also described that families expected to receive medications with every call. They were generally unaware of the specific criteria (e.g., illness, severity, and location) that determined eligibility for medication delivery.

### Impact

All staff reported that their involvement with the TMDS had a positive impact on their personal and professional lives. On a personal level, staff expressed increased satisfaction from helping their communities and benefitting from higher income. Nurses gained specific skills, including experience with the principles of telemedicine, pediatric care, dosing medication, technology applications, and awareness of self-education methods (e.g., referencing guidelines). Physicians shared that they found personal satisfaction in providing more affordable and accessible healthcare to their community. They also noted that exposure to telemedicine brought an innovative dimension to their practice. Delivery drivers reported feeling greater respect and a sense of security from belonging to an institution. Professionally, drivers increased their familiarity with technology and gained a deeper geographic knowledge of their communities.

Staff reported that the TMDS had positively impacted the community as well. They emphasized that the TMDS reduces barriers (e.g., financial, security, transportation) to accessing healthcare, prevents illnesses from progressing into emergencies, reduces overspending on hospital care, creates jobs in the community, increases respect for disadvantaged people, and improves health literacy among the community. Despite the overall positive impact of the TMDS on staff and the community, there existed several challenges to the durability and scalability of the intervention. Staff safety, potential for burnout, limited scope of the TMDS model, funding, and the humanitarian and political crises in Haiti were presented by nurses and drivers as risks to the long-term sustainability of the TMDS.

## DISCUSSION

In this study, call center and delivery nurses, delivery drivers, and on-call physicians shared their perspectives on the barriers and facilitators to scaling the TMDS healthcare model through focus group sessions and written questionnaires. The areas of concentration spanned the entire workflow from virtual exams at the call center to in-person exams at the household, as well as communication systems connecting staff to each other and families. Expectations and impact were assessed to determine the extent of alignment with the broader goal of improving accesses to pre-emergent care for children. The results have enabled staff and study leadership to develop solutions aimed at refining the service in a way that anticipates scaling (Table 2). The findings can also provide direction to other teams faced with similar challenges.

**Table 2.** TMDS staff perspectives on barriers to scale and potential solutions.

| Domain | Challenges | Solutions |
| --- | --- | --- |
| Virtual exam | • Lack of confidence in the accuracy of patient details provided by parents over the phone | • Request ‘read back’ from parents to ensure two-way understanding<br>• Provide a ‘tip sheet’ with common follow-up and exploratory questions<br>• Establish a standardized pathway for handling situations when the nurse is uncertain about the quality of information<br>• Eliminate or modify unproductive components of the virtual exam |
| Call center environment | • Technological barriers (e.g., unstable cellular signal, changing user interfaces) | • Enhance the robustness of technology infrastructure and establish contingencies<br>• Strengthen and diversify technology training |
| In-person exam | • Restrictive scope of workflow and clinical resources | • Develop strategies to address barriers that prevent families from seeking emergency care<br>• Facilitate interactions with hospital access points<br>• Refine the workflow to accommodate situations in which additional patients are encountered during a household visit |
| Household environment | • Maneuvers to arrive at the household and function effectively once there | • Assure dispatch of nurses to conduct in-person exams is indicated<br>• Innovate nurse equipment to increase lighting, maximize space, and improve durability |
| Clinical resources | • Limitations on medication dosing and edge cases<br>• Informal workflow for consulting on-call physicians | • Incorporate clinical decision support tools for medication dosing<br>• Protocolize edge cases within the clinical resources<br>• Create structured systems for presenting cases and outcomes to on-call physicians |
| Communication | • Unstable cellular connectivity disrupts communication chains<br>• Unreliable patient addresses | • Create an office in the delivery zone, rather than utilize mobile workstations<br>• Focus on obtaining proper names for individuals and locations |
| Expectations | • Family expectations exceed the scope of the TMDS | • Improve messaging on the scope of a TMDS with staff and within the community |
| Impact | • Risks to long-term sustainability could limit impact | • Foster in-depth partnerships and distribute risk<br>• Ensure standards of professionalism are supported and continue offering professional development opportunities |

The process of evaluation through focus groups, written questionnaires and a framework matrix analysis inspired an unanticipated higher-ordered synthesis and visualization of the strengths and weaknesses of the TMDS model (Figure 2). This visualization divides the findings into two domains: procedural workflow and governing mission. Once the workflow is set to a virtual or in-person encounter, nurses must have immediate situational awareness and adapt accordingly. This involves a cognitive process of decision making and a series of physical actions. In parallel, the situational awareness and adaptations require communication with a spectrum of different types of people with variable access to connectivity (e.g., cellular networks). These activities are hard, stressful and carry risk. Motivation to succeed with these difficult tasks is founded in a commitment to mission. This mission extends from self to community, motivates personal and team-based creative problem-solving, and requires recognition that the service has limited scope and capacity.

**Figure 2.**
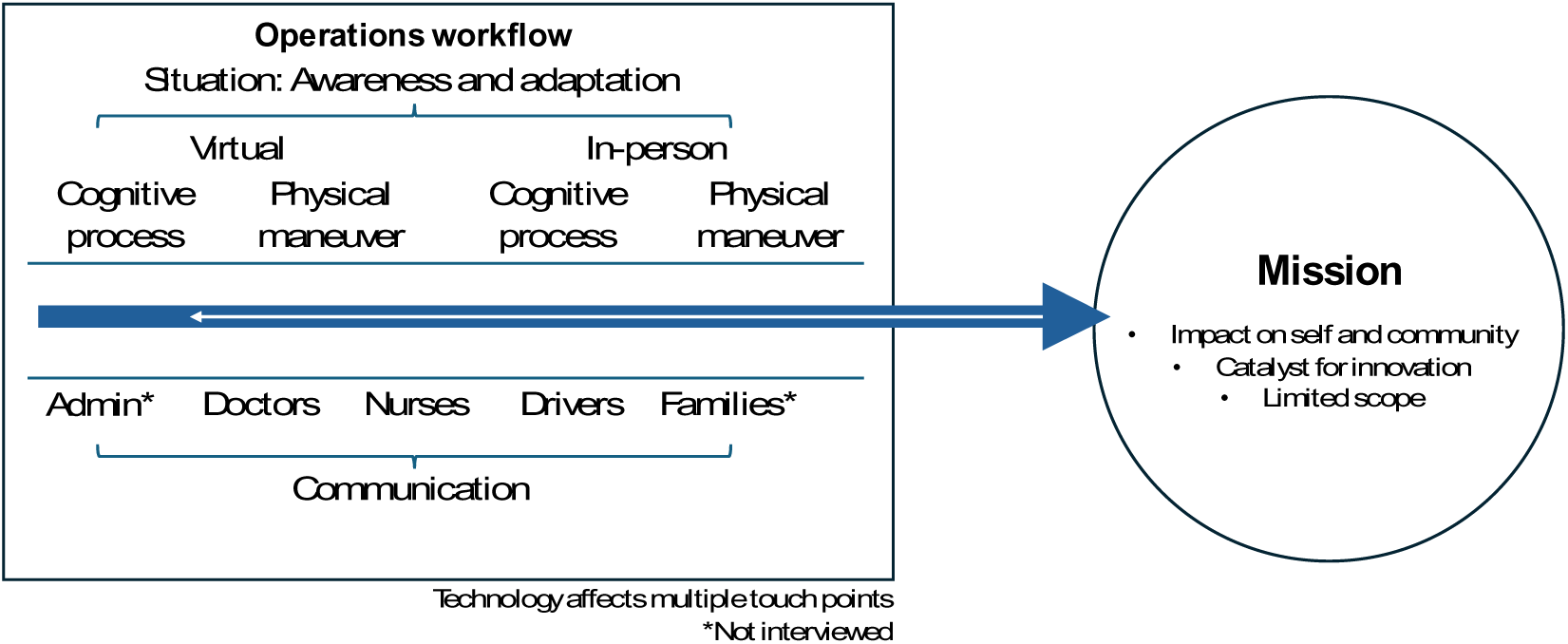
Positioning a TMDS for scale; barriers and solutions between operations and mission.

With this visualization in mind, several solutions were identified, relating to both the cognitive and physical challenges. Cognitive challenges included struggling to effectively phrase questions to conduct productive virtual exams, as well as difficulty managing situations outside the scope of the established workflow and clinical resources. While nurses mitigated these challenges through experience, the time required to gain sufficient experience poses a barrier to scaling. To address this, consolidating acquired knowledge into a ‘tip sheet’ and protocolizing how to handle edge-cases might expedite the training of new providers. Additionally, implementing an eCDS tool that digitizes the clinical guidelines may streamline onboarding and provide structured guidance for conducting exams. Finally, refining components of the virtual and in-person exam processes could further enhance the quality of treatment plans.

Solutions to address the physical challenges focused on strengthening the technology infrastructure and innovating resources. In settings like Haiti, where telecommunication services are unreliable yet critical for telemedicine, improving the telecommunication infrastructure at its source may not be feasible. However, establishing multiple contingencies at each touchpoint (‘hardening’) can help minimize communication barriers. Additionally, households where in-person exams are conducted lack the resources typically found in clinics. To improve the working conditions, innovating materials to provide adequate lighting, a designated workspace, and improved organizational capacity would strengthen the quality and efficiency of in-person encounters.

Finally, unmet expectations from both families and staff, along with risks to the long-term sustainability of the TMDS, present obstacles to the mission. During this initial pilot phase, the scope of the TMDS was intentionally limited in terms of age, clinical offerings, hours of operation, and workflow to establish proofs of concept. As the TMDS is positioned for scale, the scope may expand. Until then, aligning the scope with staff and community expectations can be achieved through clear and consistent messaging. Furthermore, an uncertain path to sustainability poses a barrier to maintaining high levels of impact and trust among both staff and community members. To mitigate these risks, increasing and diversifying partnerships, while ensuring high standards for working conditions and professional development, may improve the likelihood of long-term scalability and sustainability.

Our results align with, as well as differ from, the experiences of other telemedicine services. In the telemedicine literature, staff and end-user acceptability are cited as common barriers [39]. While we did not encounter opposition to telemedicine, misaligned expectations suggest underlying concerns related to acceptability. Another frequently cited barrier involves factors related to the implementation environment, such as nationwide policies and legal liability [28, 40]. These concerns did not apply to our controlled pilot implementation but may become relevant as the service scales. The limited technical abilities of staff were identified as a barrier in our study and is consistent with findings from other research [39, 41]. Additionally, most telemedicine services, particularly those operating in low resource settings, face persistent challenges related to unreliable telecommunication infrastructure [40, 41].

This study was not without limitations. First, data on family expectations and community impact were based solely on staff impressions, which may have introduced bias. Future research could address this limitation by directly enrolling families as participants to obtain firsthand insights. Second, the translation process could have introduced information bias, potentially altering the nuances and exact meanings of responses from the original language. To mitigate this risk we worked with bilingual facilitators and had three team members involved in translating and confirming transcriptions before analysis began. Third, conducting focus groups via video conference may have introduced a second form of information bias, as some sections of the recordings were inaudible due to unstable connectivity. Despite these limitations, the study yielded important findings with both internal value for the TMDS and broader relevance to the telemedicine research community.

## CONCLUSION

Telemedicine has secured a permanent role in healthcare delivery globally. However, scaling novel telemedicine applications presents challenges. To better understand these challenges, we used a framework matrix analysis to synthesize insights from focus group discussions and written questionnaires completed by TMDS staff. The findings highlighted key areas for improvement and led to the development of essential action items, organized into three domains; conceptual, physical and mission-oriented. These action items will guide internal efforts to position the MotoMeds TMDS for scale and will hopefully catalyze external improvements among like-minded telemedicine initiatives.

## Data Availability

All data produced in the present study are available upon reasonable request to the authors

## ADDITIONAL FILES

Additional File 1. PDF. Agenda to guide focus group discussions.

Additional File 2. PDF. Questionnaire for call center and delivery nurses (INACT3)

## Abbreviations

TMDS: telemedicine and medication delivery service
CFIR: Consolidated Framework for Implementation Research
ICT: communication technology infrastructure
INACT-H: Improving Nighttime Access to Care and Treatment-Haiti
IT: information technology
eCDS: electronic clinical decision support

## ADDITIONAL INFORMATION

### Ethics statement

This study was reviewed and approved by the University of Florida Institutional Review Board (202002693) as well as the Comité National de Bioéthique (National Bioethics Committee of Haiti; Ref2021-11). Consent process described in manuscript.

### Consent for publication

Not applicable.

### Availability of data and materials

The datasets used and/or analysed during the current study are available from the corresponding author on reasonable request.

### Code availability

Not applicable.

### Author contributions

Conceptualization: EJN, MBK, TKB, RKR,

Implementation of the study: FEG, KC, MBK

Statistical analyses and visualizations: FEG, KC, KF, EJN, MBK

Funding acquisition: MBK, EJN

Supervision: MBK, EJN, VBR, JRB, LE, CB

Writing – original draft: MBK, EJN

Writing – review & editing: RKR, FEG, KC, YC, KEF, JRB, TKB, LE, VBR, CB, MBK, EJN

All authors read and approved the final manuscript.

## Acknowledgements

We would like to thank the TMDS staff for their enrollment and participation in this study. We would also like to acknowledge the University of Florida Department of Pediatrics administrators Randy Autrey, Krista Berquist and Brittney Johnson. Additionally, we express our appreciation for the academic support provided by Glenn Morris at the Emerging Pathogens Institute, and Rashmin Savani Chair of the Department of Pediatrics.

## REFERENCES

1. Bhaskar S, Bradley S, Chattu VK, Adisesh A, Nurtazina A, Kyrykbayeva S, Sakhamuri S, Yaya S, Sunil T, Thomas P et al: Telemedicine Across the Globe-Position Paper From the COVID-19 Pandemic Health System Resilience PROGRAM (REPROGRAM) International Consortium (Part 1). Frontiers in Public Health 2020, 8.

2. Ndwabe H, Basu A, Mohammed J: Post pandemic analysis on comprehensive utilization of telehealth and telemedicine. Clinical eHealth 2024, 7:5–14.

3. Sood S, Mbarika V, Jugoo S, Dookhy R, Doarn CR, Prakash N, Merrell RC: What is telemedicine? A collection of 104 peer-reviewed perspectives and theoretical underpinnings. Telemed J E Health 2007, 13(5):573–590.

4. Telemedicine Opportunities and Developments in Member States [https://apps.who.int/iris/bitstream/handle/10665/44497/9789241564144_eng.pdf]

5. Liu P, Wang F, Xu W, Li Y, Li B: Trends and frontiers of research on telemedicine from 1971 to 2022: A scientometric and visualisation analysis. Journal of Telemedicine and Telecare 2023, 29(9):731–746.

6. Harst L, Otto L, Timpel P, Richter P, Lantzsch H, Wollschlaeger B, Winkler K, Schlieter H: An empirically sound telemedicine taxonomy – applying the CAFE methodology. Journal of Public Health 2022, 30(11):2729–2740.

7. Telehealth for emergency departments. Tele-triage [https://telehealth.hhs.gov/providers/best-practice-guides/telehealth-for-emergency-departments/tele-triage#:~:text=Tele%2Dtriage%20can%20be%20used,come%20to%20the%20emergency%20department]

8. McConnochie KM, Conners GP, Brayer AF, Goepp J, Herendeen NE, Wood NE, Thomas A, Ahn DS, Roghmann KJ: Effectiveness of Telemedicine in Replacing In-Person Evaluation for Acute Childhood Illness in Office Settings. Telemedicine and e-Health 2006, 12(3):308–316.

9. Mahmoud K, Jaramillo C, Barteit S: Telemedicine in Low- and Middle-Income Countries During the COVID-19 Pandemic: A Scoping Review. Front Public Health 2022, 10:914423.

10. Alnasser Y, Proaño A, Loock C, Chuo J, Gilman RH: Telemedicine and Pediatric Care in Rural and Remote Areas of Middle-and-Low-Income Countries: Narrative Review. J Epidemiol Glob Health 2024, 14(3):779–786.

11. Moyo J, Madziyire G: Use of telemedicine in obstetrics and gynaecology in Zimbabwe during a lockdown period. Pan Afr Med J 2020, 35(Suppl 2):89.

12. Klarman MB, Flaherty KE, Chi X, Cajusma Y, Capois AC, Vladimir Dofiné MD, Exantus L, Friesen J, Beau de Rochars VM, Baril C et al: Implementation of a pediatric telemedicine and medication delivery service in a resource-limited setting: A pilot study for clinical safety and feasibility. The Journal of Pediatrics 2022.

13. Flaherty KE, Klarman MB, Zakariah AN, Mahama MN, Osei-Ampofo M, Nelson EJ, Becker TK: Evaluating the prerequisites for adapting a paediatric nighttime telemedicine and medication delivery service to a setting with high malarial burden: A cross-sectional pre-implementation study. Trop Med Int Health 2023, 28(9):763–770.

14. McCannon CJ, Berwick DM, Massoud MR: The Science of Large-Scale Change in Global Health. JAMA 2007, 298(16):1937–1939.

15. Hanson K, Ranson MK, Oliveira-Cruz V, Mills A: Expanding access to priority health interventions: a framework for understanding the constraints to scaling-up. Journal of International Development 2003, 15(1):1.

16. Zomahoun HTV, Ben Charif A, Freitas A, Garvelink MM, Menear M, Dugas M, Adekpedjou R, Légaré F: The pitfalls of scaling up evidence-based interventions in health. Glob Health Action 2019, 12(1):1670449.

17. ExpandNet [https://expandnet.net/]

18. Bulthuis SE, Kok MC, Raven J, Dieleman MA: Factors influencing the scale-up of public health interventions in low- and middle-income countries: a qualitative systematic literature review. Health Policy Plan 2020, 35(2):219–234.

19. Damschroder LJ, Reardon CM, Widerquist MAO, Lowery J: The updated Consolidated Framework for Implementation Research based on user feedback. Implement Sci 2022, 17(1):75.

20. Huang F, Cheng S, Du X, Chen W, Scano F, Falzon D, Wang L: Electronic recording and reporting system for tuberculosis in China: experience and opportunities. J Am Med Inform Assoc 2014, 21(5):938–941.

21. Ndlovu K, Scott RE, Mars M: Interoperability opportunities and challenges in linking mhealth applications and eRecord systems: Botswana as an exemplar. BMC Medical Informatics and Decision Making 2021, 21(1):246.

22. Spicer N, Berhanu D, Bhattacharya D, Tilley-Gyado RD, Gautham M, Schellenberg J, Tamire-Woldemariam A, Umar N, Wickremasinghe D: ‘The stars seem aligned’: a qualitative study to understand the effects of context on scale-up of maternal and newborn health innovations in Ethiopia, India and Nigeria. Globalization and Health 2016, 12(1):75.

23. Adebayo PB, Oluwole OJ, Taiwo FT: COVID-19 and Teleneurology in Sub-Saharan Africa: Leveraging the Current Exigency. Frontiers in Public Health 2021, 8.

24. Mengiste SA, Antypas K, Johannessen MR, Klein J, Kazemi G: eHealth policy framework in Low and Lower Middle-Income Countries; a PRISMA systematic review and analysis. BMC Health Services Research 2023, 23(1):328.

25. Pérez-Noboa B, Soledispa-Carrasco A, Padilla VS, Velasquez W: Teleconsultation Apps in the COVID-19 Pandemic: The Case of Guayaquil City, Ecuador. IEEE Engineering Management Review 2021, 49(1):27–37.

26. Jiang Y, Sun P, Chen Z, Guo J, Wang S, Liu F, Li J: Patients’ and healthcare providers’ perceptions and experiences of telehealth use and online health information use in chronic disease management for older patients with chronic obstructive pulmonary disease: a qualitative study. BMC Geriatrics 2022, 22(1):9.

27. Aydemir S, Ocak S, Saygılı S, Hopurcuoğlu D, Haşlak F, Kıykım E, Aktuğlu Zeybek Ç, Celkan T, Demirgan EB, Kasapçopur Ö et al: Telemedicine Applications in a Tertiary Pediatric Hospital in Turkey During COVID-19 Pandemic. Telemedicine and e-Health 2020, 27(10):1180–1187.

28. Dodoo JE, Al-Samarraie H, Alzahrani AI: Telemedicine use in Sub-Saharan Africa: Barriers and policy recommendations for Covid-19 and beyond. International Journal of Medical Informatics 2021, 151:104467.

29. Hamilton AB, Finley EP: Qualitative methods in implementation research: An introduction. Psychiatry Res 2019, 280:112516.

30. Proctor E, Silmere H, Raghavan R, Hovmand P, Aarons G, Bunger A, Griffey R, Hensley M: Outcomes for Implementation Research: Conceptual Distinctions, Measurement Challenges, and Research Agenda. Administration and Policy in Mental Health and Mental Health Services Research 2011, 38(2):65–76.

31. Busetto L, Wick W, Gumbinger C: How to use and assess qualitative research methods. Neurological Research and Practice 2020, 2(1):14.

32. Rosen RK, Gainey M, Nasrin S, Garbern SC, Lantini R, Elshabassi N, Sultana S, Hasnin T, Alam NH, Nelson EJ et al: Use of Framework Matrix and Thematic Coding Methods in Qualitative Analysis for mHealth: The FluidCalc app. International Journal of Qualitative Methods 2023, 22:16094069231184123.

33. Klarman M, Schon J, Cajusma Y, Maples S, Beau de Rochars VEM, Baril C, Nelson EJ: Opportunities to catalyse improved healthcare access in pluralistic systems: a cross-sectional study in Haiti. BMJ Open 2021, 11(11):e047367.

34. Klarman MB, Chi X, Cajusma Y, Flaherty KE, Capois AC, Dofiné MDV, Exantus L, Friesen J, Beau de Rochars VM, Becker T et al: Development and evaluation of a clinical guideline for a paediatric telemedicine service in a low-resource setting. BMJ Paediatr Open 2024, 8(1).

35. Klarman MB, Chi X, Cajusma Y, Flaherty KE, Beausejour JR, Exantus L, Beau de Rochars VM, Baril C, Becker TK, Gurka MJ et al: Evaluation of a Scalable Design for a Pediatric Telemedicine and Medication Delivery Service: A Prospective Cohort Study in Haiti. medRxiv 2024:2024.2008.2016.24312128.

36. Flaherty KE, Klarman MB, Cajusma Y, Schon J, Exantus L, Beau de Rochars VM, Baril C, Becker TK, Nelson EJ: A Nighttime Telemedicine and Medication Delivery Service to Avert Pediatric Emergencies in Haiti: An Exploratory Cost-Effectiveness Analysis. The American Journal of Tropical Medicine and Hygiene 2022, 106(4):1063–1071.

37. Biswas D, Hossin R, Rahman M, Bardosh KL, Watt MH, Zion MI, Sujon H, Rashid MM, Salimuzzaman M, Flora MS et al: An ethnographic exploration of diarrheal disease management in public hospitals in Bangladesh: From problems to solutions. Social Science & Medicine 2020, 260:113185.

38. Rosen RK, Garbern SC, Gainey M, Lantini R, Nasrin S, Nelson EJ, Elshabassi N, Alam NH, Sultana S, Hasnin T et al: Designing a Novel Clinician Decision Support Tool for the Management of Acute Diarrhea in Bangladesh: Formative Qualitative Study. JMIR Hum Factors 2022, 9(1):e33325.

39. Scott Kruse C, Karem P, Shifflett K, Vegi L, Ravi K, Brooks M: Evaluating barriers to adopting telemedicine worldwide: A systematic review. J Telemed Telecare 2018, 24(1):4–12.

40. Mamuye A, Nigatu AM, Chanyalew MA, Amor LB, Loukil S, Moyo C, Quarshie S, Antypas K, Tilahun B: Facilitators and Barriers to the Sustainability of eHealth Solutions in Low- and Middle-Income Countries: Descriptive Exploratory Study. JMIR Form Res 2023, 7:e41487.

41. Sagaro GG, Battineni G, Amenta F: Barriers to Sustainable Telemedicine Implementation in Ethiopia: A Systematic Review. Telemedicine Reports 2020, 1(1):8–15.

