## Additional File 1 for "Pathways to optimize a pediatric telemedicine and medication delivery service: A multi-level qualitative study in Haiti"

**Protocol Title: "**Pathways to optimize a pediatric telemedicine and medication delivery service: A qualitative study from nurse, physician and driver perspectives"

**Version:** April 27, 2022

**Study title:** Novel approach to improve patient care and diarrheal disease research using mobile technology
**Subtitle:** Improving Nighttime Access to Care and Treatment (PART 3); INACT3

**Focus groups to identify opportunities to improve MotoMeds' clinical and logistical procedures.**

**Objectives:** From a nurse provider perspective, identify opportunities to improve the clinical and logistical workflows, patient interactions (call center and household), guideline adherence, and case report form for both paper and digital iterations. From a delivery driver perspective, identify opportunities to improve logistical workflows and interactions with families. From an on-call physician perspective, gather feedback on the role and how well it integrates within the larger workflow. From the perspective of all team members, consider to what extent MotoMeds meets or does not meet the expectations of families, as well as how the team members' involvement with MotoMeds has had a positive impact at the personal, professional, and community levels.

**Methods:**
Consent. Two bilingual and impartial facilitations will complete waivers of documentation of consent and data collection. They will train by completing/reviewing UF relevant trainings for human subjects research and FHI 360's Qualitative Research Methods: A Data Collector's Field Guide.

Nurse providers. Eight active call center nurses (Gressier) and 6 active on-call nurses (Les Cayes) will be offered participation. Nurses who agree to participate will be given a written questionnaire (see Nurse Questionnaire) to be completed and returned during a work shift. The completed questionnaires will be linked to the respondents by study IDs. The facilitators will review the surveys for completeness and follow up with the individual nurses if further clarification is needed. Additionally, all nurses will be offered participation in focus groups that will be conducted virtually in groups of 3-4 nurses each and follow the agenda below.
Delivery drivers. Seven active delivery drivers in Gressier and 6 active delivery drivers in Les Cayes will be offered participation. Drivers who agree to participate will be given a short biographical form (see below) to fill out prior to beginning the focus group discussion. Facilitators will conduct the focus groups virtually and each group will have 3-4 drivers and follow the agenda below.
On-call physicians. Three active on-call physicians will be offered participation. Physicians who agree to participate will be given a short biographical form (see below) to fill out prior to beginning the focus group discussion. Facilitators will conduct the focus group virtually.

Focus groups. The focus groups will be conducted by 2 facilitators (when possible) in Haitian Kreyol and the audio/video will be recorded. The facilitators will transcribe the recordings into English for data analysis. A debrief will be completed by each facilitator following every focus group. The debrief will provide facilitators with an opportunity to reflect on how the focus groups went, if they were receiving relevant responses and if there are any minor changes needed prior to continuing. The debrief will also provide study leadership with an opportunity to monitor progress.

**Nurses**

1. **Introductions (5 mins)**
   Intent- *The objective of part 1 is to introduce the facilitators and explain the purpose of the focus groups. We are looking to gather ideas regarding opportunities to improve clinical and logistical workflows of the call center and household interactions. Additionally, we want to consider to what extent MotoMeds meets or does not meet the expectations of families, as well as how the team members' involvement with MotoMeds has had a positive impact at the personal, professional, and community levels.*
   a) Thank you for participating, we will be introducing some topics to discuss, the conversation will take about 1 hour.
   b) Introductions
   c) We will record the discussion and then making a transcript and an English translation of this discussion. When we do this, we remove your name and other things that identify you from the responses. We keep your comments confidential and the recording of this interview will be kept on a secure server and will be deleted at the end of the study.

2. **Clinical successes and challenges with call center interactions (15 mins)** Gressier nurses only
   *Intent- The objective of part 2 is to learn about the clinical successes and challenges nurse providers face when consulting patients at the call center and solicit suggestions for improvement.*
   a) In what situations are you confident in the information you are receiving over the phone during consultations?
   b) In what situations are you not confident in the information you are receiving over the phone during consultations?
      i. What do you do in these situations?
   c) Have you encountered a clinical situation that you did not know how to manage at the call center? Explain.
      i. Are there any tools that may have better helped you manage the situation?
      Facilitators explain/show what a digital clinical decision support tool might look like. Use demo of Outbreak Responder Dehydration Assessment Calculator
   d) Do you think a digital version of the case report form embedded with clinical decision support would be useful?
      i. In what ways?
   e) How could the role of the on-call physician be improved?

3. **Logistical challenges at the call center (10 mins)** (Gressier nurses only)
*Intent- The objective of part 3 is to learn about the logistical challenges nurse providers face when consulting patients and dispatching drivers at the call center and solicit suggestions for improvement. Facilitators should review challenges from the nurse questionnaire and can reference specific challenges during the discussion.*
   a) What do you do when you run into challenges with technology at the call center?

b) What ideas do you have to overcome challenges communicating with the Les Cayes nurses?
c) What ideas do you have to overcome challenges dispatching drivers and/or on- call nurses in Gressier?

**4. Clinical and logistical challenges with household interactions (10 mins)**
*Intent- The objective of part 4 is to learn about the clinical and logistical challenges nurse providers face when consulting patients at the household and offer suggestions for improvement. We encourage you to provide feedback even if you do not currently perform household visits.*
a) Have you encountered a clinical situation that you did not know how to manage at the household? Are there tools that may have better helped you manage the situation?
b) Have you encountered a technical situation that you did not know how to manage at the household? Are there tools that may have better helped you manage the situation?

**5. Working from home (10 mins)** Les Cayes and Gressier on-call nurses only
*Intent- The objective of part 6 is to learn about the challenges nurse providers face when working from their home and solicit suggestions for improvement.*
a) With respect to the workflow of being on call at home and preparing for a delivery/household visit, what challenges do you face? How can these challenges be solved?
b) Are there any essential resources that are lacking to work out of your home? What is needed?

**6. User expectations and improvements (10 mins)**
*Intent- The objective of part 7 is to get your impression of how well MotoMeds meets the expectation of families who use the service. Think about comments that families have made, both positive and negative at any point in the call center or household interaction. You do not need to focus only on the questions about feedback during the 10-day follow up call. We also want to hear your ideas how we could improve the service for families.*
a) Do you have ideas of how the service could be improved?
b) What do you think are the biggest challenges to MotoMeds sustainability?

**7. Impact and professional development (15 mins)**
*Intent- The objective of part 5 is to better understand how MotoMeds has impacted the drivers personally. This could be on a professional level through training and increased income or more abstract from serving their community.*
a) How has MotoMeds positively impacted you? Your family? Your community?
b) What skills have you gained during your experience with MotoMeds?

**Delivery drivers**

1. **Introductions (5 mins)**
   Intent- *The objective of part 1 is to introduce the facilitators and explain the purpose of the focus groups. We are looking to identify opportunities to improve logistical workflows and interactions with families. Additionally, we want to consider to what extent MotoMeds meets or does not meet the expectations of families, as well as how the team members' involvement with MotoMeds has had a positive impact at the personal, professional, and community levels.*
   a) Thank you for participating, we will be introducing some topics to discuss, the conversation will take about 30 minutes.
   b) Introductions
   c) We will audio record the discussion We will be making a transcript and an English translation of this discussion. When we do that, we remove your name and other things that identify you. We keep your comments confidential and the audio recording of this interview is kept on a secure server and will be deleted at the end of the study.

2. **Technical (15 mins)**
   *Intent- The objective of part 2 is to learn about the challenges drivers face when making deliveries or transporting nurses.*
   a) What challenges do you face in communication with nurses?
   b) What challenges do you face in communication with other drivers?
   c) What challenges do you face in communication with families?
   d) How might we improve communication in general?
   e) What are the strengths of using dispatch software (Beacon)?
   f) What are the weaknesses of using dispatch software (Beacon)?
   g) Beyond communication, how might we improve workflow (e.g., length of videos)?

3. **Family interaction (15 mins)**
   *Intent- The objective of part 3 is to learn more about interactions between drivers and families.*
   a) Do families ask you questions beyond the scope of your responsibilities? How do you deal with that?
   b) If someone in the community were to describe what MotoMeds is to their neighbor/friend what would they say?

4. **User expectations (15 mins)**
   *Intent- The objective of part 4 is to get your impression of how well MotoMeds meets the expectation of families who use the service. Think about comments that families have made, both positive and negative at any point in interacting with them over the phone or in person.*
   a) What comments do families make about the service when you are completing deliveries?
   b) What expectation do families have that MotoMeds accomplishes most often?
   c) What expectation do families have that MotoMeds fails to accomplish?
   d) Do you have ideas for how MotoMeds could be improved?
   e) What do you think are the biggest challenges to the sustainability of MotoMeds?

5. **Impact and professional development (15 mins)**

<u>Intent</u>- *The objective of part 5 is to better understand how MotoMeds has impacted the drivers personally. This could be on a professional level through training and increased income or more abstract from serving their community.*

c) How has MotoMeds positively impacted you? Your family? Your community?
d) What skills have you gained during your experience with MotoMeds?

**On-call physicians**

1. **Introductions (5 mins)**
   <u>Intent</u>- *The objective of part 1 is to introduce the facilitators and explain the purpose of the focus groups. We are looking to better understand your perspective of the effectiveness of the on-call physician role and how well it integrates within the larger workflow. Additionally, we want to consider to what extent MotoMeds meets or does not meet the expectations of families, as well as how the team members' involvement with MotoMeds has had a positive impact at the personal, professional, and community levels.*
   a) Thank you for participating, we will be introducing some topics to discuss, the conversation will take about 30 minutes.
   b) Introductions
   c) We will record the discussion. We will be making a transcript and an English translation of this discussion. When we do that, we remove your name and other things that identify you. We keep your comments confidential and the audio recording of this interview is kept on a secure server and will be deleted at the end of the study.

2. **Clinical (15 mins)**
   <u>Intent</u>- *The objective of part 2 is to get feedback from the physicians about how comfortable they are with their clinical responsibilities. They have a difficult role to play when; they are receiving information third hand on cases that are either complicated or out of the ordinary.*
   a) In what ways are some cases presented to you appropriate for pre-emergency telemedicine?
   b) In what ways are some cases presented to you not appropriate for pre-emergency telemedicine?
   c) Do you feel you receive sufficient information about the cases to provide clinical advice?
   d) How might the nurses better present cases to you?
   e) Do you have concerns about effectively conveying the physician's plan to nurses? Explain.

3. **Workflow (15 mins)**
   <u>Intent</u>- *The objective of part 3 is to get feedback from the physicians about how their role fits in within the larger MotoMeds workflow.*
   a) Do you think the consult role of the on-call physician is effective? If not, what improvements can be made?
   b) Do you think the oversight role of the on-call physician is effective? If not, what improvements can be made?
   c) Do you feel connected to the project and other team members? If not, what improvements can be made?
   d) How might the workflow be improved for the on-call physician?

4. **Impact and professional development (15 mins)**
   <u>Intent-</u> *The objective of part 4 is to better understand how MotoMeds has impacted the on-call physicians personally. This could be on a professional level through gaining experience or more abstract from serving their community.*
   a) How has MotoMeds positively impacted you? Your family? The community?
   b) Has your professional development been impacted by your position with MotoMeds?

**Biographical information**

- Age
- Sex
- Phone number
- Years of experience working in your profession
- Years/months of experience in current position with MotoMeds
- Highest level of school/training completed
