## Additional File 2 for "Pathways to optimize a pediatric telemedicine and medication delivery service: A multi-level qualitative study in Haiti"

Questionnaire for call center nurses (INACT3)

*Biographical*

a) Study ID:

b) What sex are you?

c) What is your age? :

d) What is your telephone number?

e) How many years of experience do you have working in your profession?

f) How many months or years do you have in your position with MotoMeds?

g) What level of school or education did you finish?

h) Are you an employer of Gressier or Les Cayes?

i) Do you work on-call?

*Informational*

1. What question in the Case Report Form is more useful when you are providing consultation on the telephone?
2. What question in the Case Report Form is not useful when you are providing consultation on the telephone?
3. Are there any questions on the Case Report Form that you would change? What changes would you have made?
4. Are there any questions that are not on the CRF that you ask regularly? Do you think we should add those questions to the CRF?
5. Is there anything in the clinical guide that you find difficult to to navigate? If so, what?
6. What problems do you run into when prescribing medications?
7. What works well in the call center?
8. What doesn't work too well in the call center?
9. What thing or question takes more time when providing consultation on the telephone?

10. Do you run into technological problems? If so, what problems?
11. What problems do you face when communicating with nurses from Les Cayes?
12. What problems do you experience in regards to dispatching drivers?
13. What problems do you experience when you nurses to visit the children’s household in Gressier?
14. What method of advertisement do you think is not successful?
15. Do you have any additional ideas for advertising?
16. What aspect in a household visit has the most importance when you are consulting a child?
17. What thing is most clinically difficult during household visit examinations?
18. What thing is most logically difficult during household visit examinations?
19. What aspect of the household visit takes the most time to complete?
20. If someone in the community were to describe MotoMeds to a neighbor, what would they say?
21. What expectations do family members have that MotoMeds often accomplishes?
22. What expectations do family members have that MotoMeds often does not accomplish?
23. How has MotoMeds positively impacted you? Your family?
24. How has MotoMeds positively impacted the community?
25. How has MotoMeds affected your professional development?
26. What competence has you gained during your experience with MotoMeds?

Questionnaire for delivery nurses (INACT3)

*Biographical*

a) Study ID:

b) What sex are you?

c) What is your age? :

d) What is your telephone number?

e) How many years of experience do you have working in your profession?

f) How many months or years do you have in your position with MotoMeds?

g) What level of school or education did you finish?

h) Are you an employer of Gressier or Les Cayes?

i) Do you work on-call?

*Informational*

1. What method of advertisement do you think is more successful?
2. Do you have any additional ideas for advertising?
3. What aspect in a household visit has the most importance when you are consulting a child?
4. What thing is most clinically difficult during household visit examinations?
5. What thing is most logically difficult during household visit examinations?
6. What aspect of the household visit takes the most time to complete?
7. What advantage do you have when working from home?
8. What disadvantages do you have when working from home?
9. If someone in the community were to describe MotoMeds to a neighbor, what would they say?
10. What expectations do family members have that MotoMeds often accomplishes?
11. What expectations do family members have that MotoMeds often does not accomplish?
12. How has MotoMeds positively impacted you? Your family?
13. How has MotoMeds positively impacted the community?

14. How has MotoMeds affected your professional development?

15. What competence have you gained during your experience with MotoMeds?
